# Cardiothoracic Ratio (CTR) Among Patients Presenting for Chest X-ray in Radiology Department at Mulago National Referral Hospital: A Patients’ Health Indicator for Clinical Application

**DOI:** 10.1101/2024.08.28.24312720

**Authors:** Alen Musisi, Rebecca Nakatudde, Oliver Namuwonge, Deborah Babirye, Ismail Kintu, Francis Olweny, Richard Malumba, Victoria Nakalanzi, Aloysius Gonzaga Mubuuke

**Author notes:** **Corresponding Author:** Alen Musisi, **E-mil:**.

## Abstract

**Introduction/background:** The heart is vital, and even minor dysfunctions can significantly impact the body. Cardiologists need always to determine heart size, which varies with physiological changes. Advanced measurement techniques are costly and often inaccessible to a common man. Measuring the cardiothoracic ratio (CTR) via conventional X-ray is a common and more affordable option, but there’s a need for even cheaper alternatives

**Objective:** To determine relationship between CTR and presenting clinical indications and to relate CTR to the body parameters to find an appropriate relationship that can be utilized in low resource facilities in determining heart size.

**Methodology:** This cross-sectional study involved 386 patients undergoing chest radiographs at Mulago National Specialized Hospital’s radiology department. Data were summarized using frequencies and percentages. Associations between the cardiothoracic ratio (CTR) and independent variables were analyzed using Pearson’s chi-square, Fisher’s exact test, Spearman’s correlation coefficient, simple linear regression, and multivariate regression. Statistical significance was set at a p-value of < 0.05.

**Results:** The median cardiothoracic ratio (CTR) was 0.46, with an interquartile range of 0.42 to 0.50. Female patients had a higher CTR than males. Significant positive correlations were found between CTR; and BMI (p < 0.001, correlation 0.21), and BSA (p = 0.016, correlation 0.12), and BSI (p < 0.001, correlation 0.19). The diagnostic accuracy of a linear regression equation containing BSA as an estimator of CTR showed relatively fair performance compared to the linear regression equations with BSI and BMI. It showed sensitivity, specificity, and positive and negative predictive values of 29.2%, 86.0%, 63.6%, and 59.0% for males, and 8.3%, 98.1%, 75.0%, and 60.7% for females, respectively.

**Conclusion:** BSA shows a moderately good relationship with CTR, while the influence of body habitus on CTR is minimal. Thus, using body parameters to predict CTR should be approached cautiously. We recommend conducting a similar study on a more diverse general population

## INTRODUCTION

The heart is vital, with small dysfunctions causing significant effects. It measures 12cm by 8.5cm by 6cm, and weighs 280-340g in males and 230-280g in females(1). Cardiomegaly is heart enlargement, evident when the heart’s width exceeds half the chest’s width on a posterior-anterior (PA) chest radiograph, indicating a pathological process(2). Determining heart size is crucial for cardiologists due to its variability based on sex, age, height, and weight. Establishing normal size ranges is essential, as slight changes may indicate pathology. What is normal for some may indicate disease in others. (3).

The cardiothoracic ratio (CTR) on a PA projection radiograph measures the ratio of heart size to chest width. It’s calculated as the maximal horizontal cardiac width divided by the maximal horizontal inner thoracic cage diameter. A CTR > 0.5 suggests cardiac enlargement(4, 5). An increased CTR on Chest X-ray is associated with an increased rate of morbidity in middle-aged and elderly groups and has a linear association with increased heart size and coronary heart disease(6). While chest radiography is cost-effective for detecting cardiomegaly and measuring CTR, computed tomography (CT), magnetic resonance imaging (MRI), and echocardiography offer more precise but expensive options to a common man(7). CTR can be influenced by factors like pregnancy, ascites, or chest deformities, leading to variability in measurements unrelated to heart conditions. Studies highlight discrepancies in CTR accuracy compared to cardiac MRI(4, 8). Research also shows varying correlations between CTR and body mass index (BMI), body surface area (BSA), and body surface index (BSI), with obesity and age impacting CTR differently(9, 10). Age has been identified as a more reliable indicator, showing an increase in CTR with advancing age(11). Despite existing research on heart size and CTR in other populations, its applicability to Uganda remains unverified.

The CTR in Ugandans remains undetermined, with limited evidence on its relationship with body parameters in this population. Moreover, low-resource settings like Uganda face challenges in accessing affordable and accurate CTR measurements. Screening currently relies on costly and accessible methods like cardiac echocardiography, MRI, and CT. For most clinicians in Uganda, the practical option for heart size assessment remains the cardiothoracic ratio using conventional x-ray. Although body size influences cardiac cavity dimensions, indexed values like BMI, BSI, and BSA are not widely used to estimate CTR.

This research aimed to optimize the use of body parameters to estimate CTR when conventional x-ray measurement is unavailable. This technique was expected to assist cardiologists and clinicians in enhancing their diagnostic capabilities without depending on expensive methods for patients. CTR determination is a valuable parameter in patient assessment in hospital emergency departments and intensive care units, aiding in risk stratification, treatment evaluation, and prognosis across various diseases, beyond cardiology. Therefore, this study sought to explore the relationship between the CTR and various clinical indications presented by patients and to examine the association between CTR and key body measurements, including BMI, BSA, and BSI.

## METHODS

### Study Design and Setting

The study used a hospital-based cross-sectional design at Mulago National Referral Hospital in the radiology department. The equipment used was a digital conventional Philips X-ray machine (model: SRO33100ROT380, serial number: 989000086482), processing 20 to 30 chest X-rays daily. This hospital serves patients referred from all regions of the country, ensuring the data collected is nationally representative

### Study Population

The study included all individuals aged 18 years and older who underwent a Chest X-ray at Mulago National Referral Hospital’s radiology department from 03^rd^ July to 29^th^ September, 2023, selected through consecutive sampling. Patients that never had request forms and critically ill patients unable to provide consent were excluded, along with those whose clinical request forms indicated heart disease or were inadequately filled or those whose chest x-rays had motion artifacts.

### Sample Size estimation

The formula for calculating sample size while using correlation coefficient in cross section study (12), 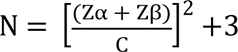, where 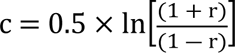, was used to estimate the number of participants needed to determine the relationship between CTR and clinical indication or CTR and body parameters. Pearson’s correlation coefficient, r=0.33 was used for clinical indications (13) and r of 0.283 was used for body mass index. To estimate the number of participants needed to describe the proportion of participants with abnormal CXR, we used Kish Leslie formula- 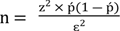 while factoring in proportion of 0.5 (ṕ) for infinite sample size because proportion of Ugandans with abnormal CTR is unknown. Margin of error of 0.05 (ԑ^2^) and Z score of 1.96 that corresponds to 95% level of confidence. We therefore came up with an overall sample size of 385 participants.

### Study Variables

The independent variables in this study were clinical indications such as cough, difficulty in breathing (DIB), and chest pain, along with demographic factors like age, height, weight, BMI, BSA, BSI, and gender. The dependent variable was CTR.

### Study Procedure

The project coordinator oversaw the data collection process conducted by research assistants, under the supervision of the Principal and Co-Principal Investigators (PI and Co-PI). Patients presenting with request forms for chest X-rays were eligible for enrollment. Research assistants provided ready written consent forms to enrolled patients for reading, they explained the study’s significance, ethical considerations, and objectives. The participants consented to the study by signing the consent form. Upon obtaining signed informed consent, demographic data such as age and clinical indications were extracted from the patients’ request forms. If necessary, missing information was obtained directly from patients by research assistants or the principal investigator. Patient weight and height were measured to calculate Body Mass Index (BMI), Body Shape Index (BSI), and Body Surface Area (BSA).

Next, the patient underwent a chest X-ray procedure using posterior-anterior (PA) projection with full inspiration. A Research Assistant obtained a soft copy of the patient’s X-ray image and securely stored it on a password-protected research computer. All measurements and relevant data for each patient were recorded on a data collection form. Subsequently, this data was entered into a data entry screen created with Epi-data software version 3.1

### Calculating CTR, BMI, BSA, and BSI

PA radiographs were conducted with the patient in an upright position, achieving maximum inspiration, and utilizing a tube-detector distance of 180cm. The Radiant DICOM software was employed to measure the transverse cardiac diameter (TCD), calculated by summing the horizontal distances from the heart’s right (a) and left (b) lateral borders to the midline (spinous processes of the vertebral bodies).

The widest transverse thoracic diameter (TTD) (c) was determined by measuring the broadest distance between the medial aspects of the ribs, specifically from pleura to pleura.

The CTR was obtained by dividing the TCD by the maximum TTD.

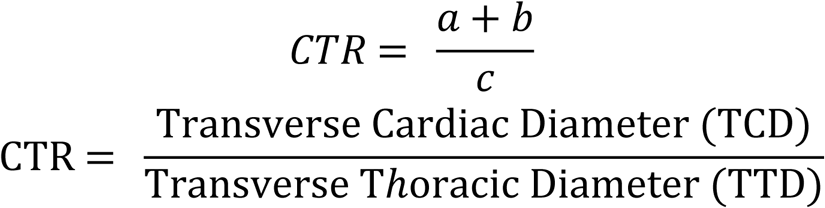

### Image demonstrating how CTR was measured

BMI was estimated mathematically as a ratio of the patient’s weight in kilograms (kg) to the square of the patient’s height in meters (m) (14);

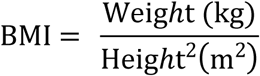

BSA was estimated from the most widely used technique called Du Bois and Du Bois formula (15);

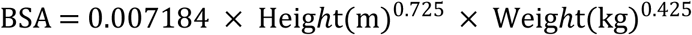

BSI, different studies show it is a more precise indicator of organ size/volume than both BMI and the BSA (16). It was estimated by dividing the both weight by the calculated square root of its BSA, it is mathematically expressed as;

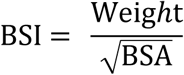

### Data Management and Analysis

Patient data was recorded in data collection forms, which were subsequently reviewed for consistency by the PI before the end of the business day. The validated data was then inputted into a password-protected data entry interface created with Epi-data software version 3.1. A backup of the entered data was stored on an external hard drive.

Data was summarized using frequencies and percentages. The relationship between CTR and independent variables was analyzed using simple linear regression and Spearman’s correlation coefficient. Significance testing was conducted using Pearson’s chi-square and Fisher’s exact test, with a p-value < 0.05 considered statistically significant.

The dataset was then divided into two parts and analyzed in two stages. The first half was stratified by gender, where multivariate linear regression was employed in each stratum. This analysis accounted for participants’ age and individually measured body parameters to establish a linear equation relating these variables to CTR.

The second half of the dataset utilized the linear equations derived from the first half to predict/estimate CTR values. This estimation considered measured body parameters and participant characteristics such as age. Subsequently, both the estimated CTR obtained from input of participants age and corresponding body parameter into the derived linear equation and the CTR determined by the study radiologist were categorized as normal (CTR value between 0.42-0.50) or abnormal (below 0.42 or above 0.50). Sensitivity, specificity, and positive and negative predictive values of the estimated CTR were then calculated against the standard of practice CTR determined by the study radiologist to diagnose abnormal heart size (considering both micro-cardia and cardiomegaly).

### Ethical Consideration

The study included patients referred for X-ray services during routine care, utilizing anonymized data comprising images and clinical information that cannot be linked back to individual patients. Ethical approval was obtained from the Makerere University, School of Medicine Research Ethics Committee (Mak-SoMREC-2022-480) at Makerere University, Kampala, Uganda. Prior to commencing the study, all investigators completed an online human subjects course.

## RESULTS

### Study profile

Out of 390 patients initially recruited for the study, 386 were ultimately enrolled. The exclusions included two patients without clinician request forms, one patient with an unclear indication for chest radiograph, and one critically ill patient unable to provide consent.

### Characteristics of the Study Participants

In the study, 386 enrolled patients included 215 females (55.7%). The median age was 37 years (interquartile range: 27-50 years), with the largest age group being 31-40 years old (27.7%). The most common clinical indication for chest radiographs was chest pain, reported by 120 patients (31.2%). These details are presented in Table 1.

**Table 1:** Characteristics of 386 patients who presented for chest radiographs in the radiology department of Mulago National Referral Hospital during the study.

| Characteristics | Frequency | Percentages |
| --- | --- | --- |
| **Gender** |  |  |
| Female | 215 | 55.7 |
| Male | 171 | 44.3 |
| **Age categories** |  |  |
| ≤ 20 | 37 | 9.6 |
| 21-30 | 92 | 23.8 |
| 31-40 | 107 | 27.7 |
| 41-50 | 61 | 15.8 |
| >50 | 89 | 23.1 |
| **Indications for CXR** |  |  |
| Chest pain | 120 | 31.2 |
| DIB | 114 | 29.6 |
| Cough | 114 | 29.6 |
| Others | 37 | 9.6 |

### Relationship between CTR and Presented Clinical Indication among patients presented for chest x-ray in Radiology Department at Mulago National Referral Hospital

The median CTR was 0.46 with an interquartile range (IQR) of 0.42-0.50. Of the 386 patients, 114 (70.5%) had CTR > 0.49. Female patients had a high CTR, 70 (61.4%) compared to male patients. No statistically significant association was observed between gender and CTR (P-value, 0.144).

The indications for taking chest radiographs were not statistically significantly associated with cardiothoracic ratio (P-value, 0.190). However, a statistically significant association was observed between age categories and CTR, (P-value <0.001). This is shown in Table 2.

**Table 2:**
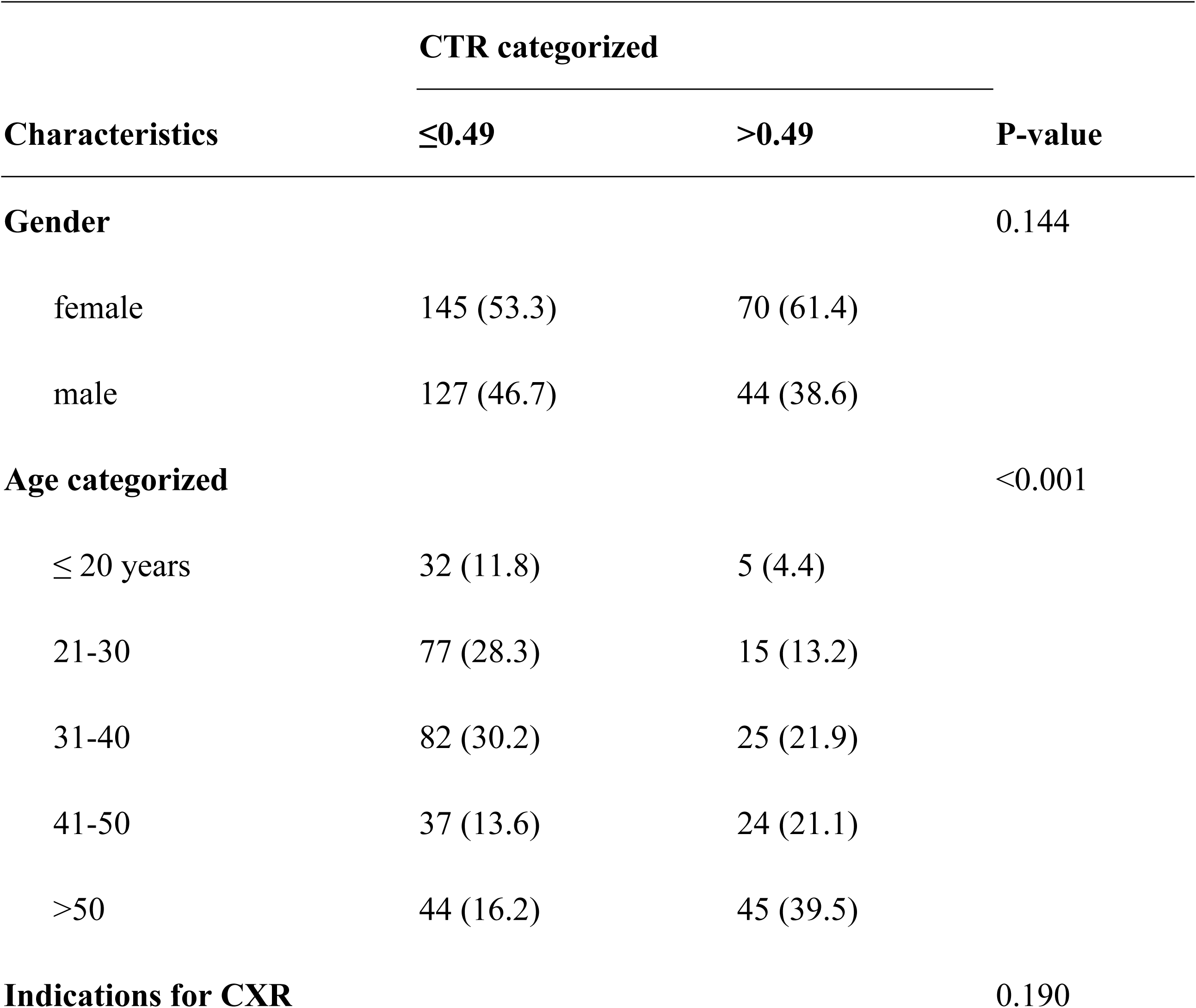
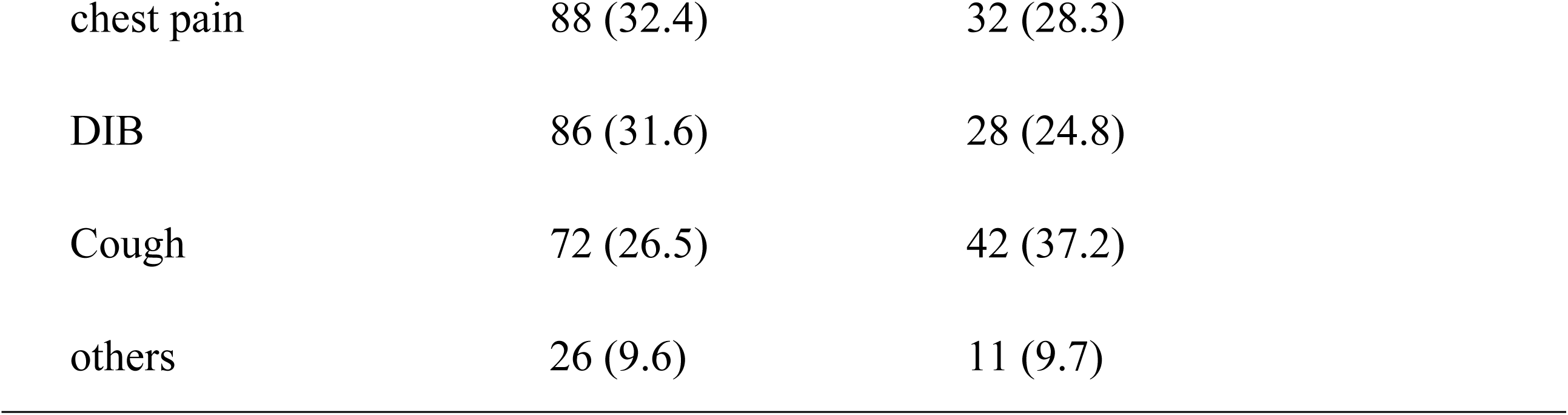
Relationship between patient characteristics and CTR among patients who presented for chest radiographs in the radiology department of Mulago National Referral Hospital during the study period.

| Characteristics | CTR categorized |  | P-value |
| --- | --- | --- | --- |
|  | ≤0.49 | >0.49 |  |
| **Gender** |  |  | 0.144 |
| female | 145 (53.3) | 70 (61.4) |  |
| male | 127 (46.7) | 44 (38.6) |  |
| **Age categorized** |  |  | <0.001 |
| ≤ 20 years | 32 (11.8) | 5 (4.4) |  |
| 21-30 | 77 (28.3) | 15 (13.2) |  |
| 31-40 | 82 (30.2) | 25 (21.9) |  |
| 41-50 | 37 (13.6) | 24 (21.1) |  |
| >50 | 44 (16.2) | 45 (39.5) |  |
| **Indications for CXR** |  |  | 0.190 |

|  |  |  |
| --- | --- | --- |
| chest pain | 88 (32.4) | 32 (28.3) |
| DIB | 86 (31.6) | 28 (24.8) |
| Cough | 72 (26.5) | 42 (37.2) |
| others | 26 (9.6) | 11 (9.7) |

### Relationship between CTR and BMI among patients presented for chest x-ray in Radiology Department at Mulago National Referral Hospital

In Table 3, it was observed that the cardiothoracic ratio (CTR) was positively correlated with BMI. This correlation was statistically significant, with a p-value of less than 0.001. However, the strength of the association was weak, as indicated by the Spearman’s rank correlation coefficient (r) of 0.21.

**Table 3:**
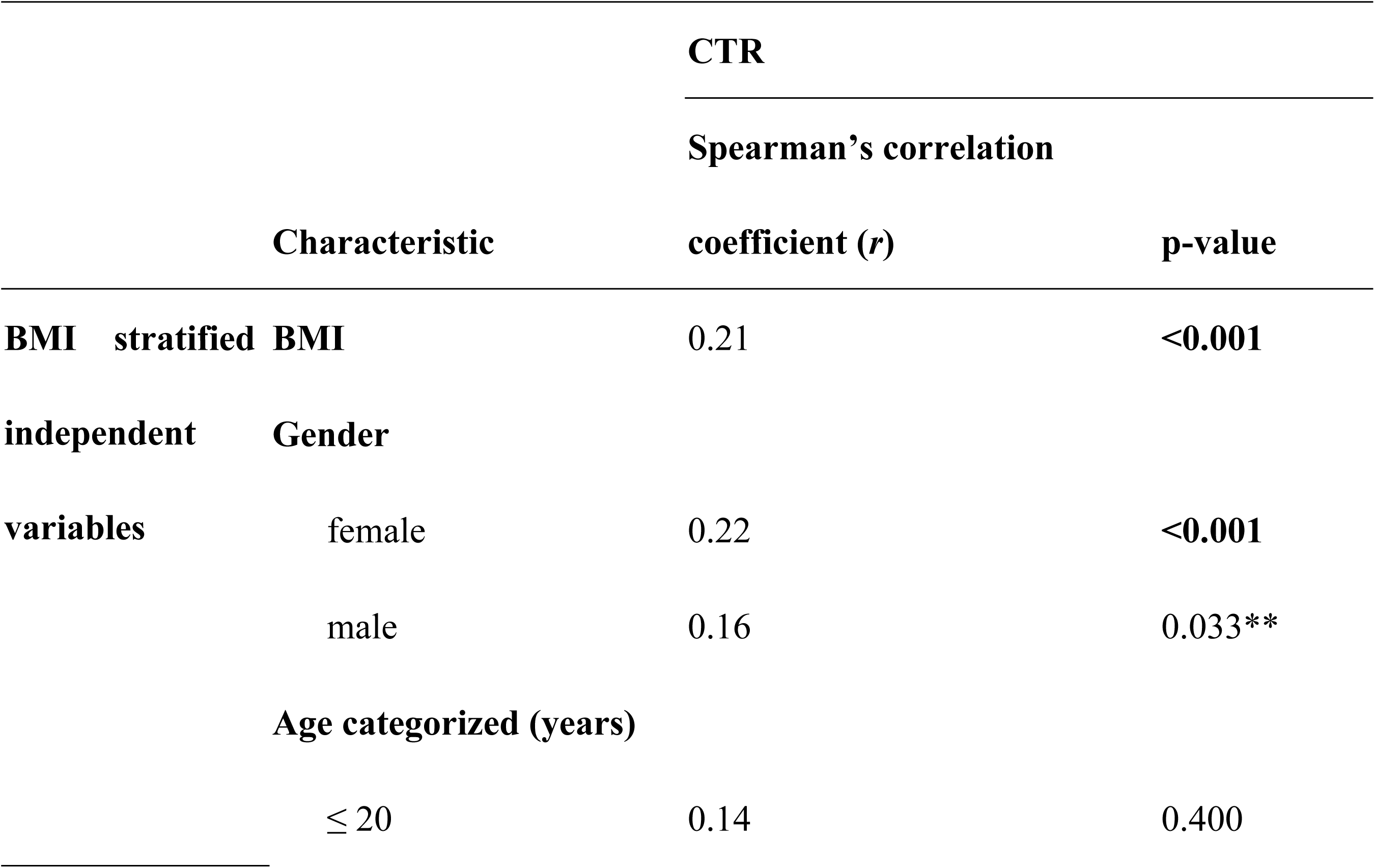
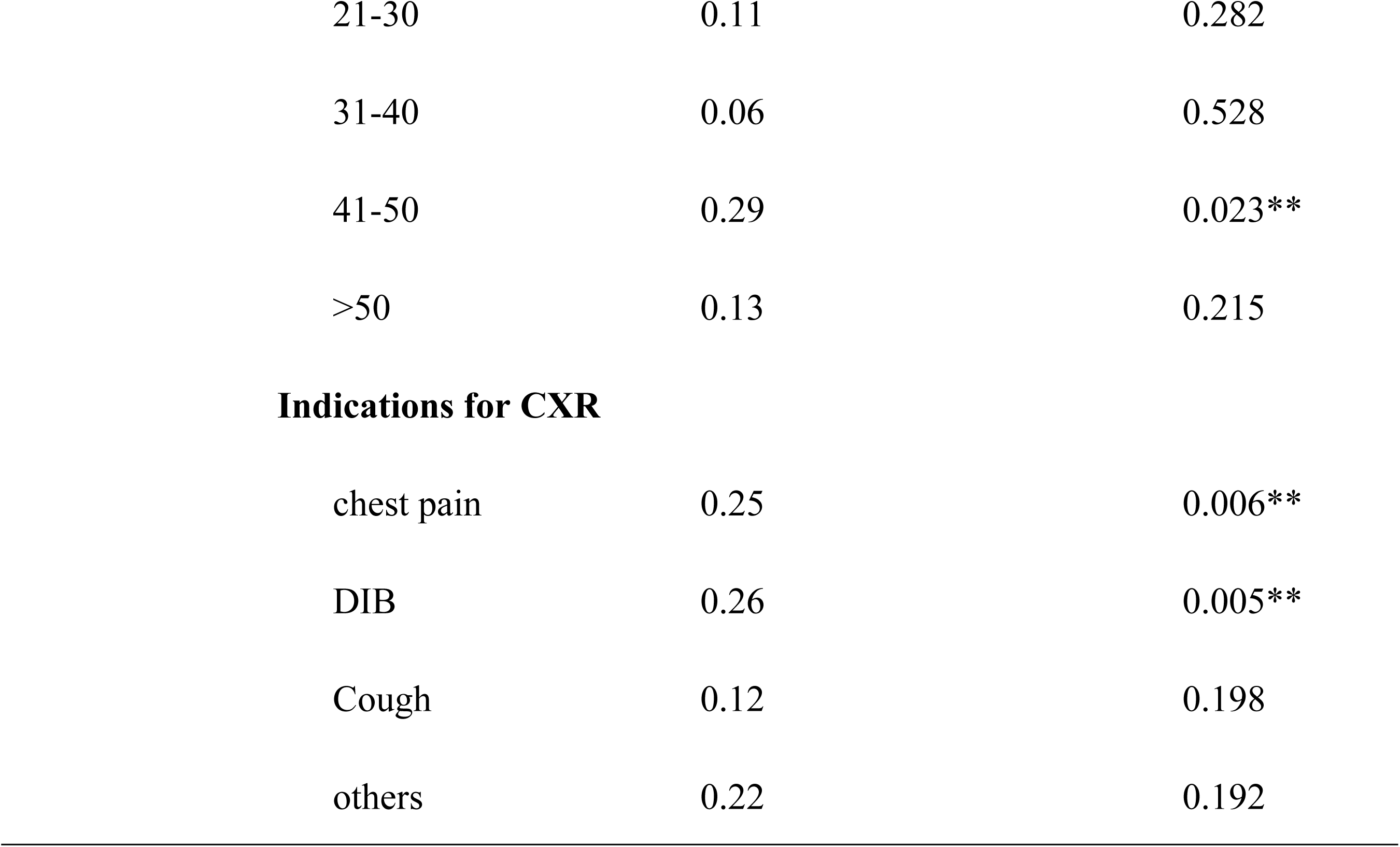
Relationship between patient BMI and CTR among patients who presented for chest radiographs stratified by gender, age, and indication at the radiology department of Mulago National Referral Hospital during the study period.

For female patients, there was a positive but weak and statistically significant correlation between BMI and CTR (r=0.22, P-value<0.001). Similarly, among male patients, there was a positive and statistically significant correlation between CTR and BMI (r=0.16, P-value, 0.033). Both of these correlations were also weak (Figure 3 and Table 3).

In Table 3, it was shown that patients aged 41-50 years had a statistically significant positive correlation between their CTR and BMI (r = 0.29, P-value = 0.023). This correlation was weak, however. BMI also showed a weak positive correlation with CTR for patients in other age categories, but these correlations were not statistically significant, with all p-values > 0.05.

The stratification of BMI based on indications for taking a chest x-ray revealed that the BMI of patients with chest pain showed a statistically significant, but weak, positive correlation with CTR (r = 0.25, P-value = 0.006). Similarly, difficulty in breathing as an indication was also found to have a statistically significant, weak positive correlation with CTR (r = 0.26, P-value = 0.005). However, there was no statistically significant correlation between BMI and CTR for patients with cough and other indications.

Figure 3 below shows that BMI for both males and females positively increased as the cardiothoracic ratio of both sexes increased however the increase was gradual and almost flat. **Graphical presentation of CTR correlation with BMI**

### 4.5 Relationship between CTR and BSA among patients presented for chest x-ray in Radiology Department at Mulago National Referral Hospital

In Table 4; the cardiothoracic ratio was positively correlated with BSA. This association was significant with a p-value of 0.016 however, the strength of the association was very weak, (r = 0.12).

**Table 4:**
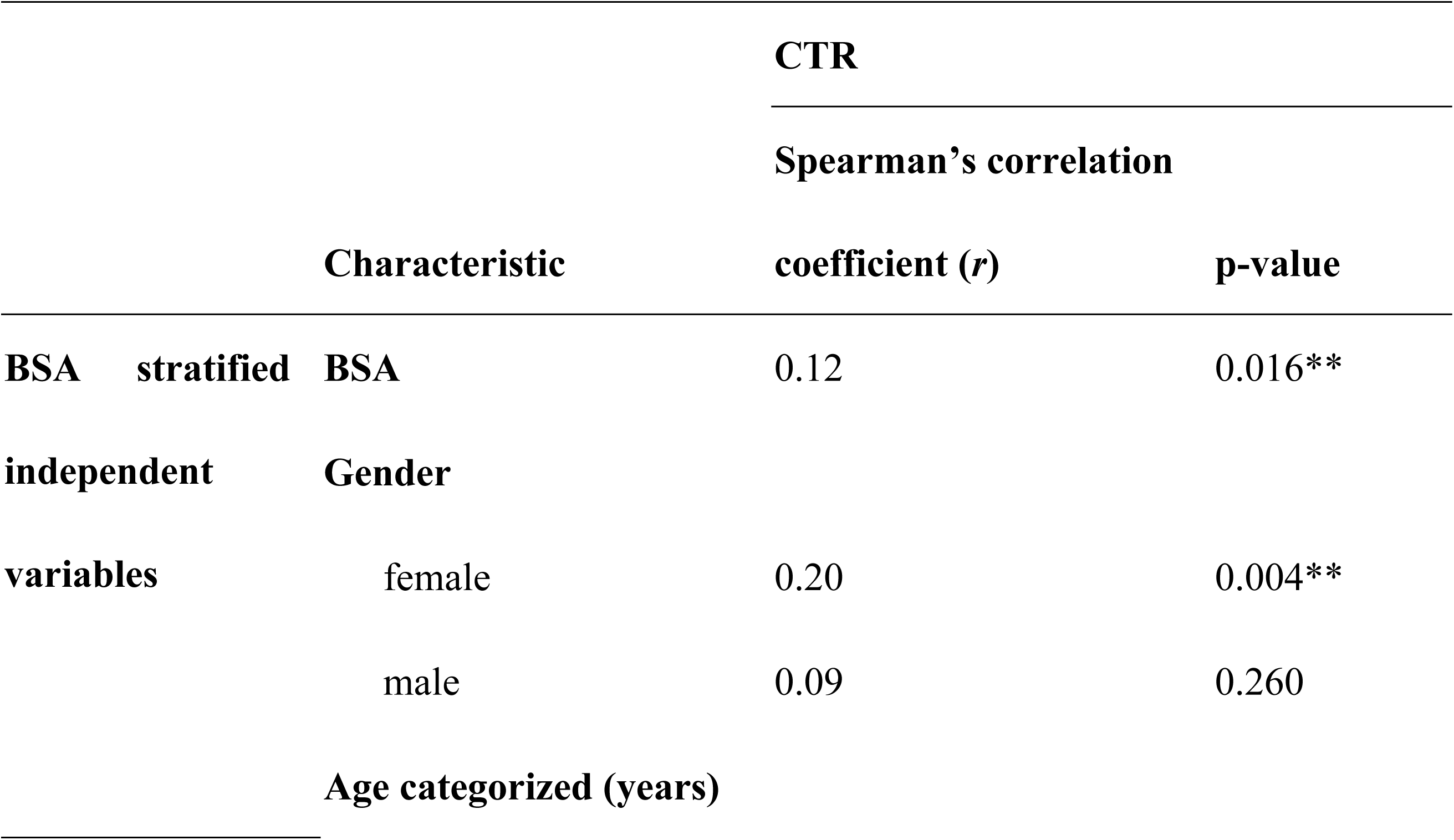
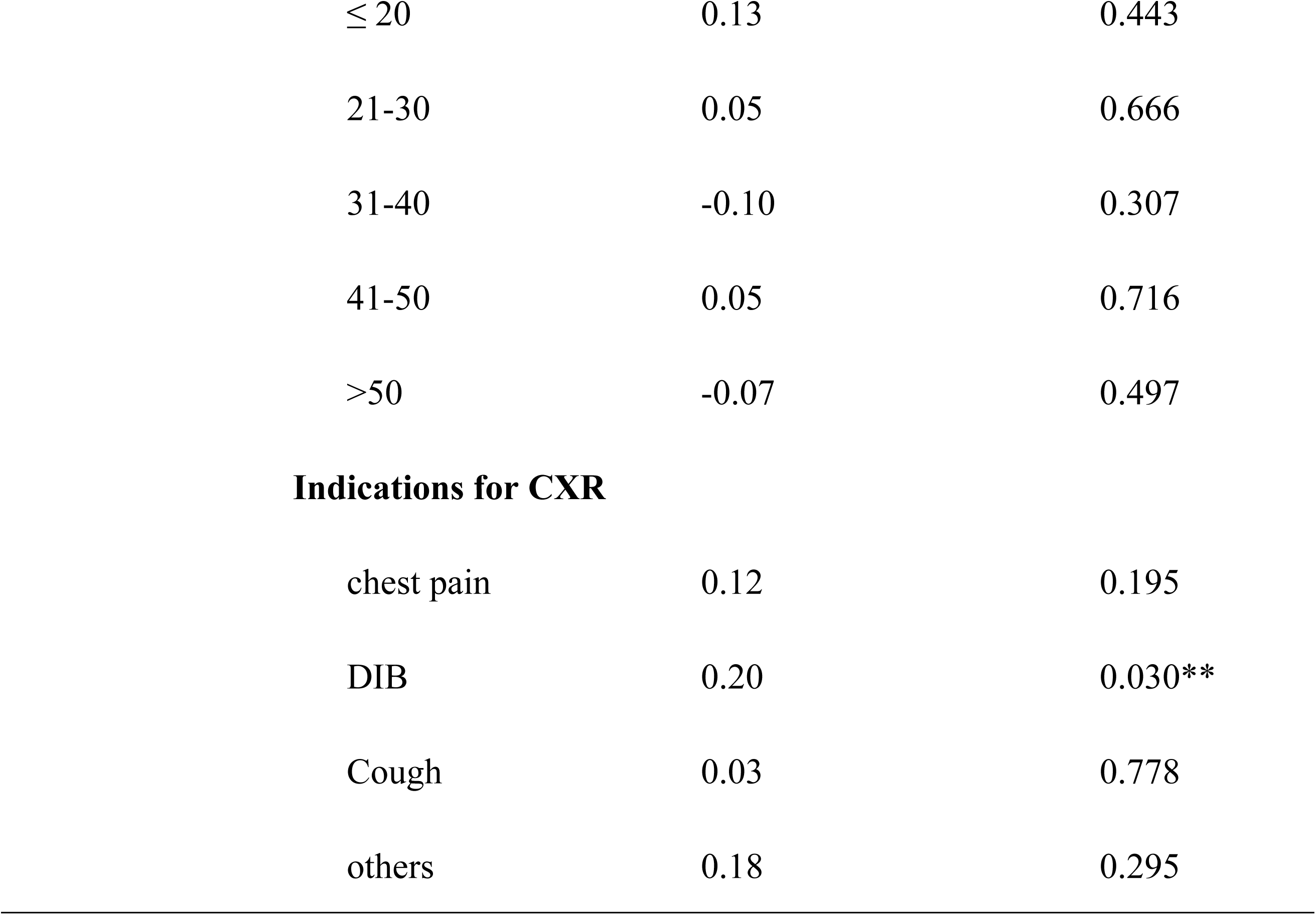
Relationship between patient BSA and CTR among patients who presented for chest radiographs stratified by gender, age and indication at radiology department of Mulago National Referral Hospital during the study period.

The correlation between BSA for female patients and their CTR was weak but positive (r = 0.20) and statistically significant (P-value = 0.004). However, the correlation between CTR and BSA among male patients though positive, was not statistically significant (P-value = 0.260) and was weak (r = 0.09). The strata of BSA by all age categories as shown in table 4 were not significantly associated with CTR. All p-values were > 0.05.

In clinical indications, DIB as strata of BSA was weak but positively correlated with CTR (r = 0.20) and was statistically significant (P-value = 0.030). Chest pain, cough, and other indications had p-values > 0.05.

Figure 4 below shows that BSA for both males and females positively increased as the cardiothoracic ratio of both sexes increased, however, the increase was gradual and almost flat.

**Graphical presentation of CTR correlation with BSA**

### 4.6 Relationship between CTR and BSI among patients presented for chest x-ray in Radiology Department at Mulago National Referral Hospital

In Table 5, the cardiothoracic ratio was positively correlated with BSI. This association was statistically significant with a p-value less than 0.001 however, the strength of the association was very weak, (*r* = 0.19). The correlation between BSI for female patients and their CTR was positive (r*=0.24*) and statistically significant (p-value < 0.001). However, the correlation between CTR and BSI among male patients though positive, was not statistically significant (p-value = 0.088) and was weak (r = 0.13).

**Table 5:**
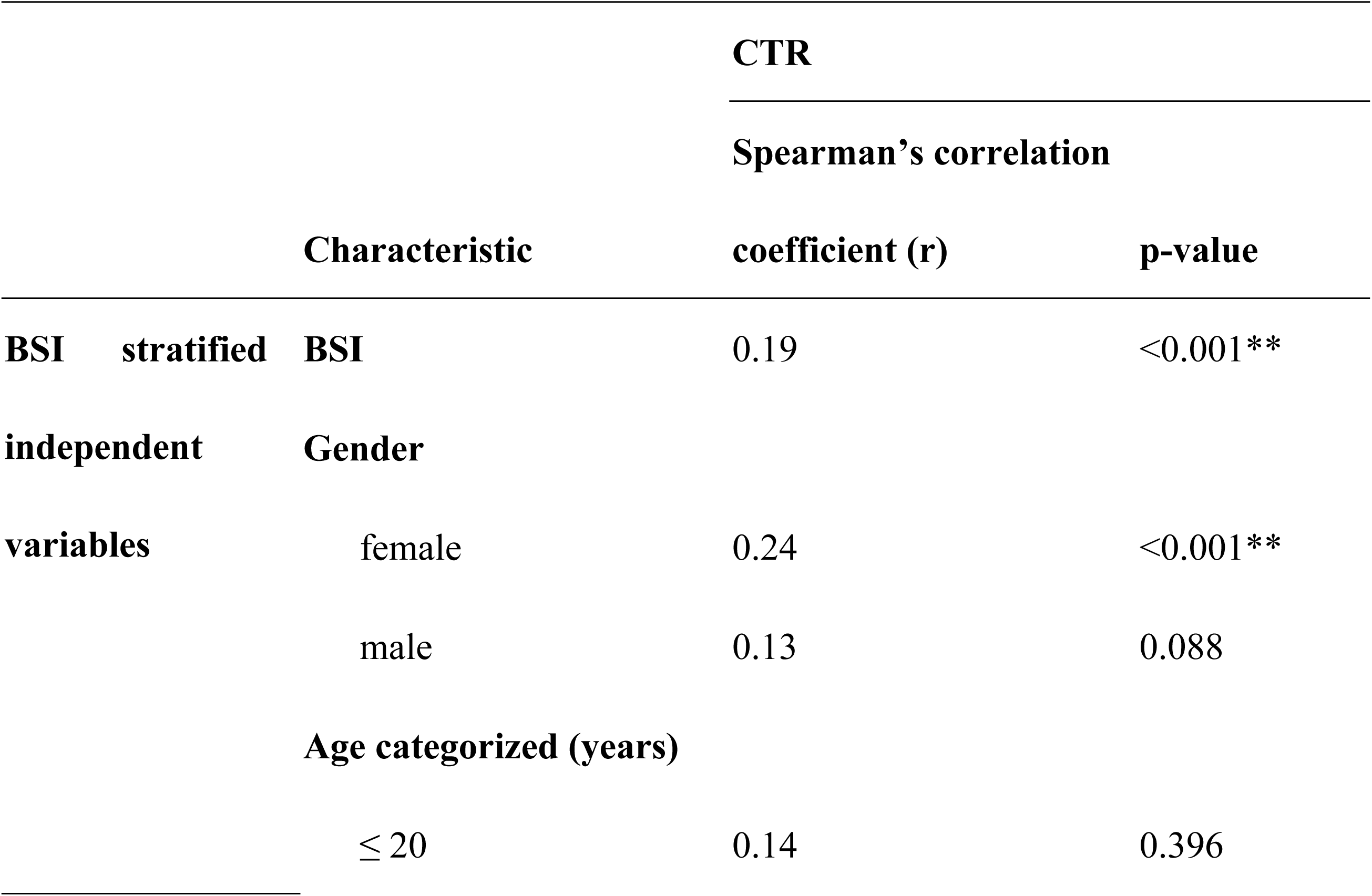
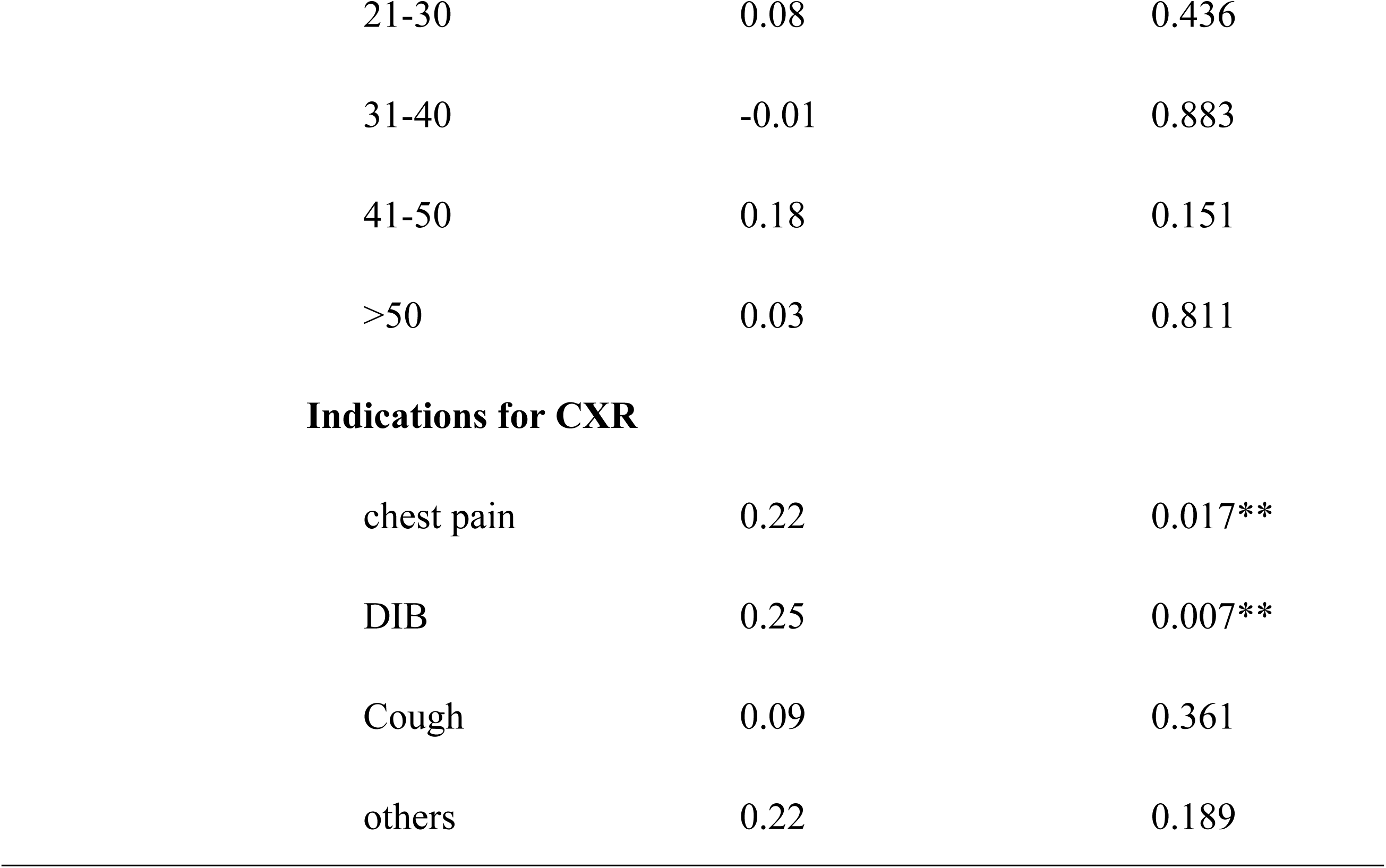
Relationship between patient BSI and CTR among patients who presented for chest radiographs stratified by gender, age and indication at radiology department of Mulago National Referral Hospital during the study period.

|  |  |  | CTR |  |
| --- | --- | --- | --- | --- |
|  |  |  | Spearman's correlation |  |
| Characteristic |  |  | coefficient (r) | p-value |
| BSI stratified independent variables | BSI |  | 0.19 | <0.001** |
|  | Gender |  |  |  |
|  |  | female | 0.24 | <0.001** |
|  |  | male | 0.13 | 0.088 |
|  | Age categorized (years) |  |  |  |
|  |  | ≤ 20 | 0.14 | 0.396 |

|  |  |  |
| --- | --- | --- |
| 21-30 | 0.08 | 0.436 |
| 31-40 | -0.01 | 0.883 |
| 41-50 | 0.18 | 0.151 |
| >50 | 0.03 | 0.811 |

Indications for CXR
|  |  |  |
| --- | --- | --- |
| chest pain | 0.22 | 0.017** |
| DIB | 0.25 | 0.007** |
| Cough | 0.09 | 0.361 |
| others | 0.22 | 0.189 |

The strata of BSI by all age categories as shown in table 5 above were all not significantly associated with CTR (p-values > 0.05). Under clinical indication, DIB as a stratum of BSI was weak but positively correlated (*r*=0.25) with CTR and statistically significant (p-value = 0.007). Chest pain as a clinical indication and stratum of BSI was also weakly but positively (*r* = 0.22) correlated with CTR and was significant (p-value = 0.017)

**Figure 4:** below shows that BSI for both males and females positively increased as the cardiothoracic ratio of both sexes increased however the increase was gradual and almost flat. **Graphical presentation of CTR correlation with BSI**

### Linear Relationship between CTR, Measured Body Parameters and Age among Male and Female Participants

In Table 6 below, it is shown that the CTR increased by about 0.0015 for every unit change in BMI while keeping age constant for female patients. CTR also increased by about 0.0017 for every unit change in age while keeping BMI constant for female patients. R-squared in this model was 0.203 meaning that about 20.3% of variations in CTR can be explained by variations in BMI and age among female patients. Even though the R-squared value was low, an F-statistic of 13.6 was significant with a p-value of less than 0.001.

**Table 6:**
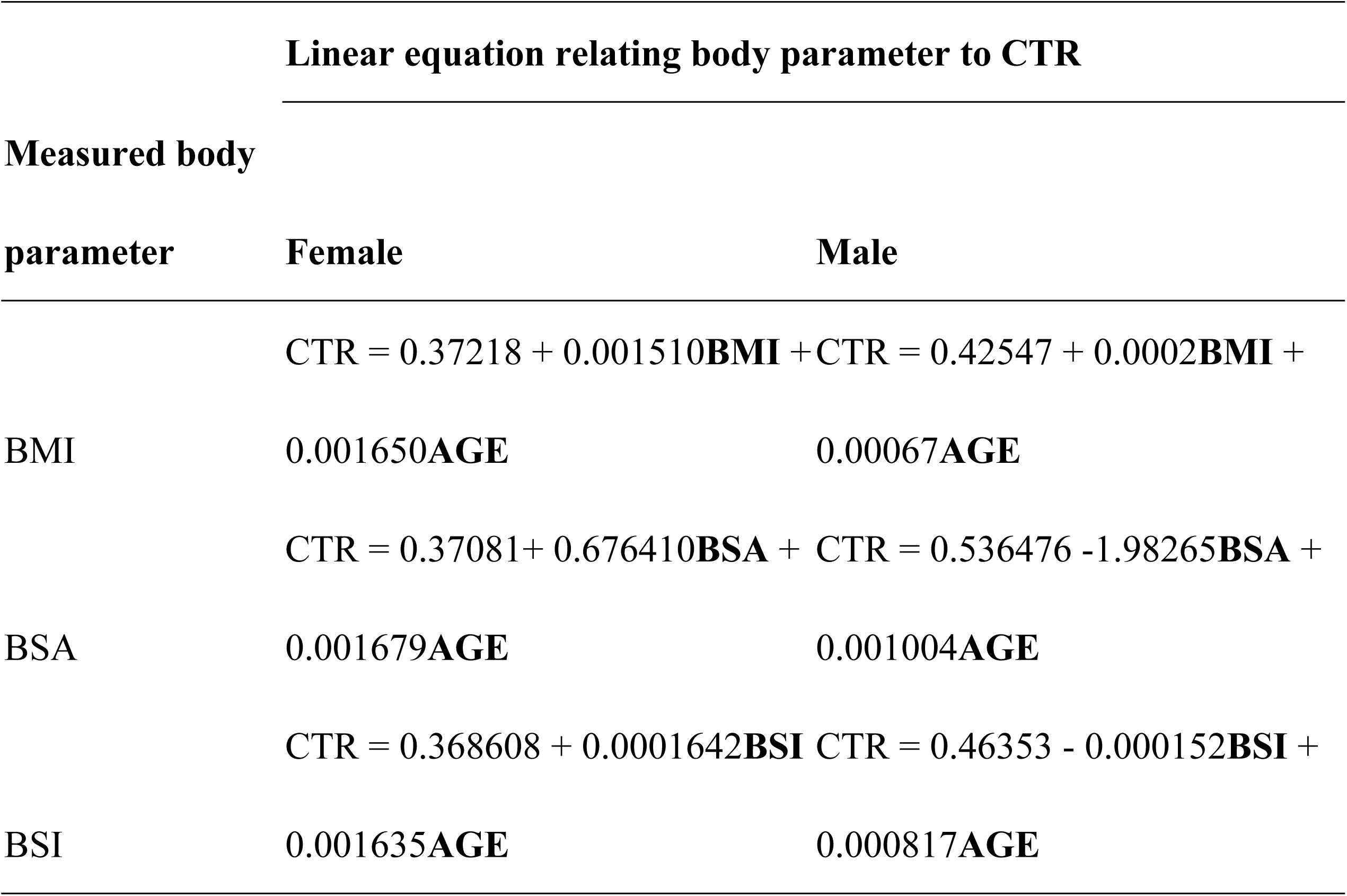
Linear relationship between measured body parameters and CTR among patients who presented for chest radiographs at radiology department of Mulago National Referral Hospital during the study period.

The study also showed that CTR increased by about 0.0002 for every unit change in BMI while keeping age constant for male patients. CTR also increased by about 0.00067 for every unit change in age while keeping BMI constant for male patients. R-squared in this modal was 0.032 meaning that only 3.2% of variation in CTR could be explained by variations in BMI and age among male patients and F-statistic of 1.33 was not significant, (0.270).

Among females, the sensitivity of estimated CTR based on the derived equation from regression of CTR against BSA and age as compared to CTR derived by the study radiologist was 29.2%, 95%CI (17.0-44.1), specificity 86.0%, 95%CI (74.2-93.7), positive and negative predictive values of 63.6%, 95%CI (40.7-82.8) and 59.0%, 95% CI (47.7-69.7) respectively.

Among males, the sensitivity of estimated CTR based on the derived equation from regression of CTR against BSA and age as compared to CTR derived by the study radiologist was 8.2%, (CI: 1.8-22.5), specificity 98.1%, (CI: 89.7-100.0), positive and negative predictive values of 75%, (CI: 19.4-99.4) and 60.7%, (CI: 49.5-71.2) respectively. This is shown in Table 7 below

**Table 7:** Sensitivity, Specificity, and Positive and Negative Predictive values for BMI, BSA, and BSI relationship with CTR among female and male patients.

| Parameters and<br>corresponding performanc<br>indicators | Female |  | Male |  |
| --- | --- | --- | --- | --- |
|  | Percentage | 95%CI | Percentage | 95%CI |
| **BMI** |  |  |  |  |
| Sensitivity | 31.3 | 18.7-46.3 | 0.0 | 0.0-9.7 |
| Specificity | 86.0 | 74.2-93.7 | 98.1 | 89.7-100.0 |
| Positive predictive value | 65.2 | 42.7-83.6 | 0.0 | 0.0-97.5 |
| Negative predictive value | 59.8 | 48.3-70.4 | 58.6 | 47.6-69.1 |
| **BSA\*** |  |  |  |  |
| Sensitivity | 29.2 | 17.0-44.1 | 8.3 | 1.8-22.5 |
| Specificity | 86.0 | 74.2-93.7 | 98.1 | 89.7-100.0 |
| Positive predictive value | 63.6 | 40.7-82.8 | 75.0 | 19.4-99.4 |
| Negative predictive value | 59.0 | 47.7-69.7 | 60.7 | 49.5-71.2 |
| **BSI** |  |  |  |  |
| Sensitivity | 31.3 | 18.7-46.3 | 0.0 | 0.0-9.7 |
| Specificity | 86.0 | 74.2-93.7 | 98.1 | 89.7-100.0 |
| Positive predictive value | 65.2 | 42.7-83.6 | 0.0 | 0.0-97.5 |
| Negative predictive value | 59.8 | 48.3-70.4 | 58.6 | 47.6-69.1 |

\*BSA was observed to have a moderate relationship with CTR and it provides a fairly better performance indicator for both males and females

## DISCUSSION

The Cardiothoracic Ratio (CTR) is a straightforward measurement and calculation, providing a practical tool as a traditional indicator of cardiac size during cardiovascular disease screenings. This study aimed to investigate the relationship between CTR and clinical indications, as well as CTR and body parameters such as BMI, BSA, and BSI. The research was conducted as a hospital-based cross-sectional study.

### Relationship between CTR and Demographics

This study found that the median CTR for both males and females was 0.46, with an IQR of 0.42-0.50. This IQR is consistent with the findings of studies by Truszkiewicz et al. and Mohammad et al., which also identified the normal CTR range as being between 0.42 and 0.50(8, 17). Several studies suggest that CTR values exceeding 0.5 or falling below 0.42 indicate pathological conditions such as cardiomegaly or microcardia, respectively (8, 17–19)

Among participants with a high CTR (>0.49), female patients outnumbered males. This finding aligns with studies by Ominde et al. and Ali et al., which attributed the higher CTR in females to their smaller thoracic diameter(18, 19). Men typically engage in more physically demanding activities, which contribute to a wider thoracic diameter(17). Unlike our study, which found no statistically significant association between gender and CTR (p=0.144), the studies by Ominde et al. and Mohammad et al. reported a statistically significant relationship(17, 18). This study also contradicted findings from a study conducted in Ghana by Brakohiapa et al., which observed a statistically significant difference in CTR across gender groups (p = 0.001) (19).

In this study, CTR was found to increase with age, a statistically significant association (p < 0.001) that aligns with findings from Brakohiapa et al., Mohammad et al., and other studies(11, 17, 19). The increase in CTR with age is attributed to the thickening of cardiac ventricular muscle due to higher vascular resistance or the loss of elasticity in the major blood vessels(11, 17). However, the combined correlation (r) between CTR and age groupings was weak, which contrasts with the Ekedigwe et al. 2014 study that found a significant correlation in males and a slightly lower correlation in females (9).

### Relationship between CTR and BMI

Our study found a statistically significant association between CTR and BMI (p < 0.001). This finding aligns with the results of the Emegoakor et al. study, which also demonstrated significant differences (p < 0.05) in CTR relative to BMI among males and females(10). In both males and females, there was a modest positive association between CTR and BMI, with correlation coefficients of 0.16 and 0.22, respectively. This finding is consistent with previous studies by Mehra et al., Ekedigwe et al., and Emegoakor et al (9–11). Similarly, this modest correlation was evident across both genders. While one might anticipate a significant correlation between CTR and BMI, our study did not demonstrate this. This discrepancy could stem from unaccounted physiological variations. Ugandans are recognized for their physical fitness, attributed to moderate-intensity work-related physical activities (20). This could lead to a slightly elevated CTR even with maintained low BMI levels. Additionally, the study only based on the clinical indications in request forms presented to the radiology department for chest x-ray and so might have missed to exclude patients with undisclosed possible cardiac conditions if not captured in the request form.

This study found a stronger statistical association between CTR and BMI in females (p < 0.001) compared to males (p = 0.033), a phenomenon not consistently reported in previous research and thus warrants further investigation. Across age categories <20, 21–30, 31–40, and >50 years, there was no statistically significant relationship between CTR and BMI, with weak positive correlations observed. The only age category where statistical significance (p = 0.023) was observed was 41–50 years, although the reason for this remains unclear.

Therefore, it’s important to note that CTR shows a weak association with BMI. BMI measurement alone does not determine CTR, as BMI calculation does not account for age, which is significantly associated with CTR.

### Relationship between CTR and BSA

Our study found a significant association (p = 0.016) between CTR and total BSA, albeit with a weak positive correlation (r = 0.12). This finding aligns with Emegoakor et al.’s study, which also reported a significant association between CTR and BSA, as well as with Mehra et al.’s study, which similarly found a weak correlation(10, 11). CTR showed statistical significance with BSA among females (p = 0.004), accompanied by a weak positive correlation. The weak correlation suggests that a twofold increase in BSA does not correspond to a twofold increase in CTR, consistent with findings from the Emegoakor et al. study(10).

A 2020 study exploring variations in cardiac geometry concerning body size among neonates with abnormal prenatal growth and birth weight indicated that adiposity influenced BSA, resulting in underestimated cardiac dimensions(21). This calls into question the use of BSA (or body weight) as a universal parameter in estimating CTR. However, a study conducted by Shiraz et al. to establish the standard reference CTR among Ghanaians demonstrated an average predictive relationship of 90.32% between CTR and BSA(22). However, the study’s limitations include the absence of statistical significance, correlation coefficients, sensitivity, and specificity values, thereby limiting its reliability.

This study identified BSA as a more effective performance indicator for both males and females, with sensitivity, specificity, positive predictive value, and negative predictive value of 29.2%, 86.0%, 63.6%, and 59.0% for males, and 8.3%, 98.1%, 75.0%, and 60.7% for females, respectively. This underscores its superiority over BMI as an indexing tool.

Furthermore, it’s notable that there was no statistical significance between CTR and BSA across different age groups, as BSA calculation does not account for age, which directly influences CTR.

### Relationship between CTR and BSI

Our study found a significant association (p < 0.001) between CTR and BSI overall, albeit with a weak positive correlation (r = 0.19). Among females, the association between CTR and BSI was also significant, with a slightly stronger correlation (r = 0.24), but less significant in males. There is limited literature on the relationship between CTR and BSI, and our study revealed a very weak relationship between the two, suggesting limited usefulness in predicting CTR.

In contrast, Shiraz et al. demonstrated that BSI provided a more accurate estimate of body size compared to BMI and BSA. Their study reported an average predictive relationship between CTR and BSI of 92.53%, higher than that observed for CTR and BSA or CTR and BMI(22). Despite this, Shiraz’s study lacked sensitivity and specificity analysis, which limits the reliability of their findings.

In this study, after incorporating age into the CTR_BSI relationship through multivariate regression, sensitivity and specificity values were observed to be more favorable for males than females, indicating that BSI may be a less effective performance indicator for predicting CTR in females.

### Relationship between CTR and Clinical Indications

In this study, we found no statistical significance between CTR and clinical indications presented (p = 0.190). This may be attributed to our study not including patients with conditions like hypertension, renal failure, and other cardiac diseases known to significantly affect CTR, as observed in studies by Hailu et al., Saheera et al., and Chou et al.(23–25) There was a statistically significant association between CTR and BMI for chest pain (p = 0.006) and difficulty in breathing (DIB) (p = 0.005), showing moderate correlations. However, there was no statistical significance observed for cough and other clinical indications in relation to CTR. Similarly, there was no statistical significance found for CTR with BSA and BSI, indicating strong correlations that support the idea that the clinical indications presented did not strongly influence CTR. This suggests that CTR is primarily affected by indications directly related to cardiac conditions.

### Diagnostic performance of linear equation containing BSA and radiologist manual method for estimation of CTR

In the current study, the performance of linear equation derived using multilinear regression method containing BSA and age showed low sensitivity of 29.2% and a relative high specificity of 86.0% in determining CTR among males. It also showed low sensitivity of 8.3% and high specificity of 98.1% among female participants. Few to no studies have used BSA and other body parameters to estimate CTR. However, studies that attempted to estimate CTR used artificial intelligence or Deep learning vs manual method of estimation by radiologists rather than body parameters. These studies have shown high sensitivity and specificity. A study by Li et al (2019) that used Deep learning computer programing showed high sensitivity of 97.2% and specificity of 92.7% in estimating CTR (26). Similarly, a study by Saiviroonporn et al (2021) showed that un assisted artificial intelligence algorithm had a standard sensitivity of 97.5% and specificity of 82.8% in estimating CTR compared (27). Clearly the difference in the findings in our study and the above studies is in the method of estimation of CTR. Deep learning and AI methods may not be reliable or might even be inaccessible, expensive and slow as the distribution of computers and access to stable constantly available electricity is still a challenge in most African countries including Uganda.

## CONCLUSION

Our study found that CTR shows minimal correlation with body parameters such as BMI, BSA, and BSI, suggesting these factors are not reliable determinants of CTR and should be used cautiously. Instead, CTR appears to be more influenced by age and sex. We observed a positive correlation between CTR and age, with higher values in females compared to males, underscoring the effectiveness of CTR measured from chest radiographs in screening for cardiomegaly in resource-limited settings.

Regarding body surface area (BSA), our study indicated a weak association with CTR. When age was included in multivariate regression analysis, BSA demonstrated relatively good performance metrics compared to BMI and BSI, with sensitivity, specificity, positive predictive value, and negative predictive value of 29.2%, 86.0%, 63.6%, and 59.0% for males, and 8.3%, 98.1%, 75.0%, and 60.7% for females, respectively. This suggests that BSA may be a more suitable tool for predicting CTR compared to BMI and BSI.

## LIMITATION(S) OF THE STUDY

These study findings may not generalize to the broader population as they were conducted in a hospital-based setting where each patient had underlying medical conditions that could have influenced their CTR.

## RECOMMENDATION(S)

We propose conducting a comparable study in a more diverse population with varied characteristics.

## Author’s contributions

AM conceptualized the idea, coordinated the project, wrote and reviewed the paper, RN was the finance director to the project, wrote and reviewed the paper, ON wrote and reviewed the paper, DB was the radiologist of the project, wrote and reviewed the paper, IS was the radiographer to the project and he reviewed the paper, FO was the data analyst to the project, wrote and reviewed the paper, RM wrote and reviewed the paper, VN wrote and reviewed the paper, AGM provided the overall mentorship of the project, wrote and reviewed the paper

## Data Availability

Data cannot be shared publicly because of ethical reasons. Data are available from the the PI (Nakatudde Rebecca) (contact via and +256788279049) for researchers who meet the criteria for access to confidential data.

## Acknowledgements

We acknowledge Makerere University Research and Innovations Fund (Mak-RIF) for funding this research. We also acknowledge Ernest Cook Ultrasound Research and Education Institute which is the primary affiliation to most of the authors including the first author.

---

**Fig 1.**
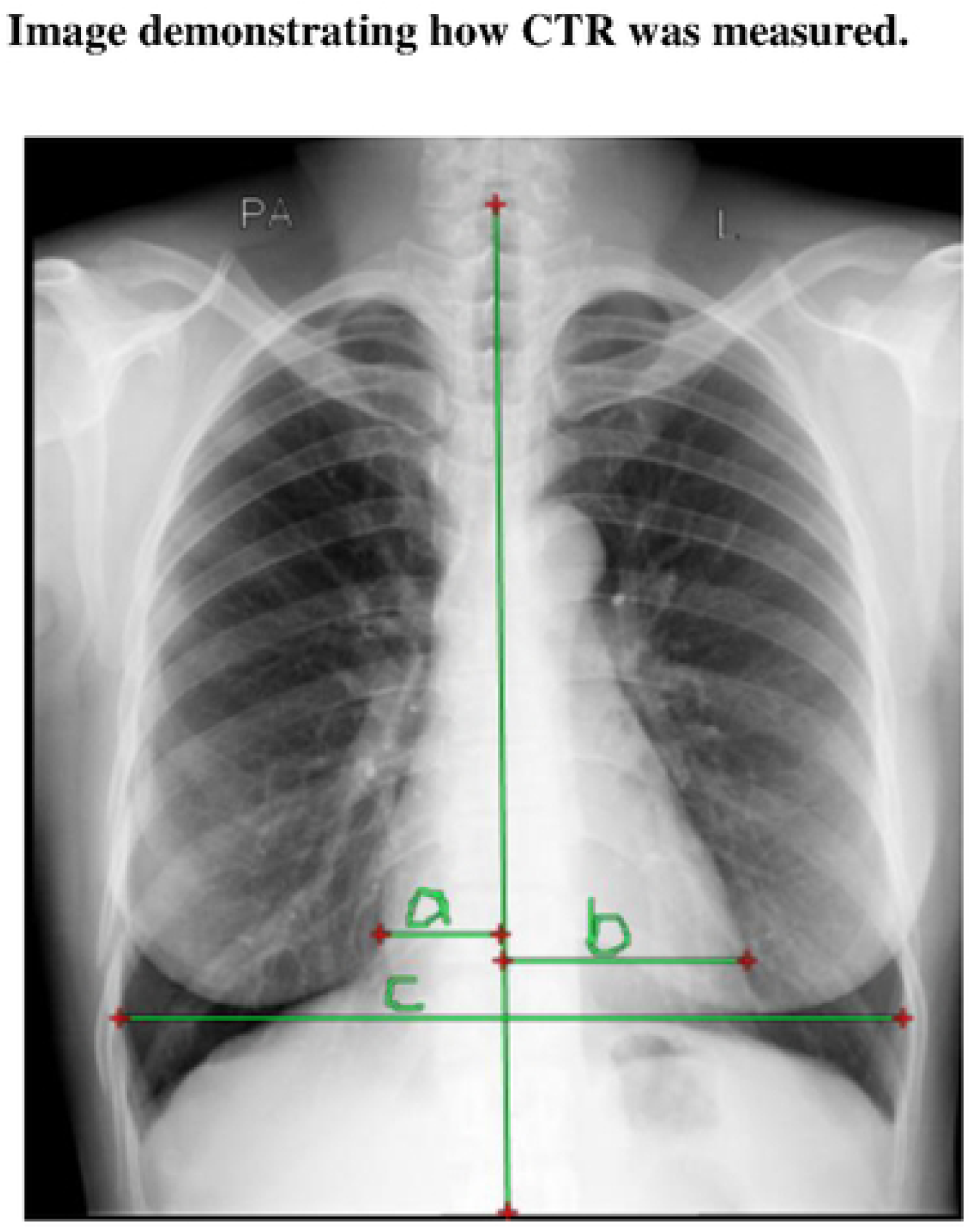
Demonstrating how CTR was measured.

**Fig 2.**
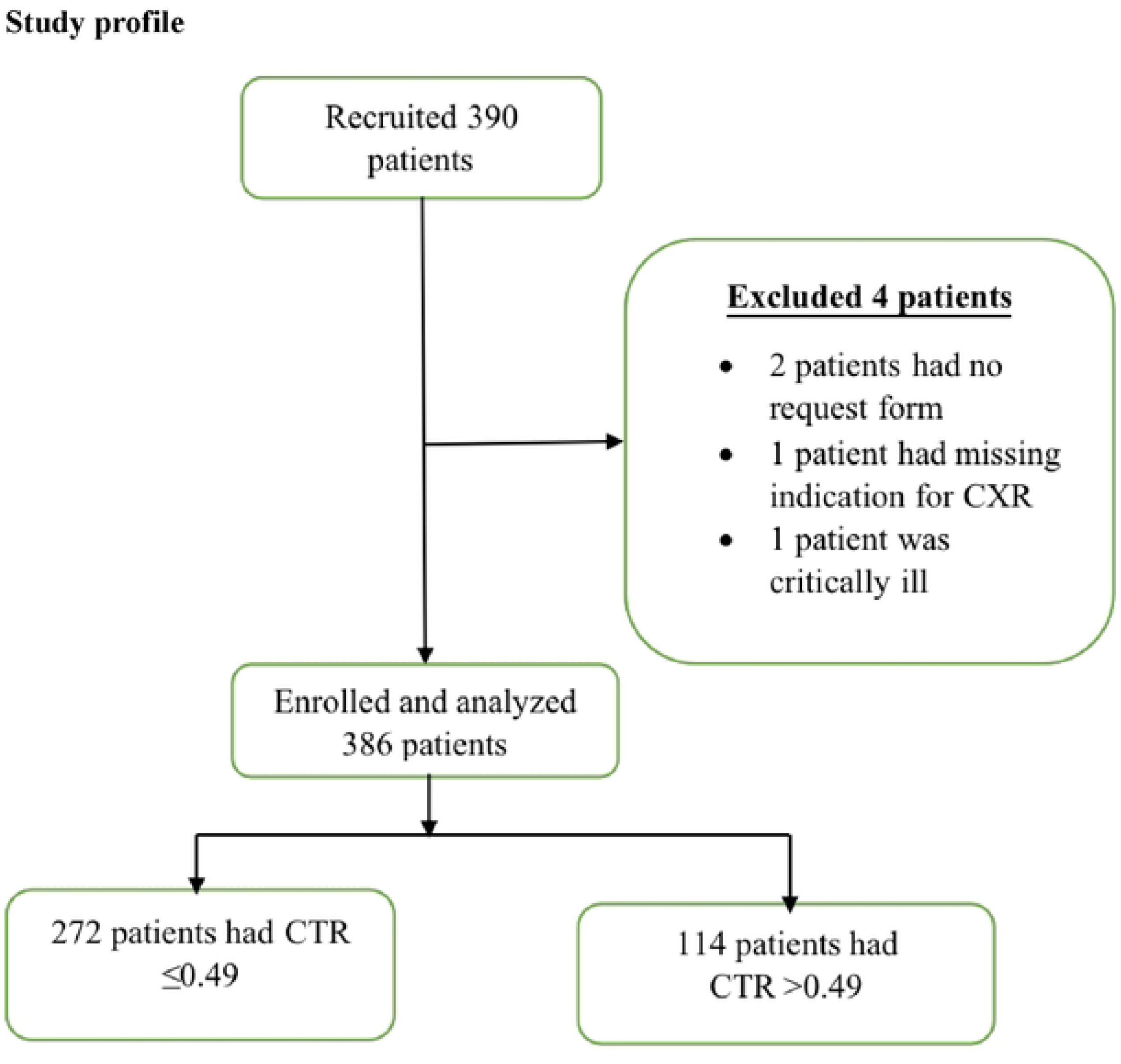
Study profile of patients presenting for chest radiographs in radiology department at Mulago National Referral Hospital during the study period.

**Fig 3.**
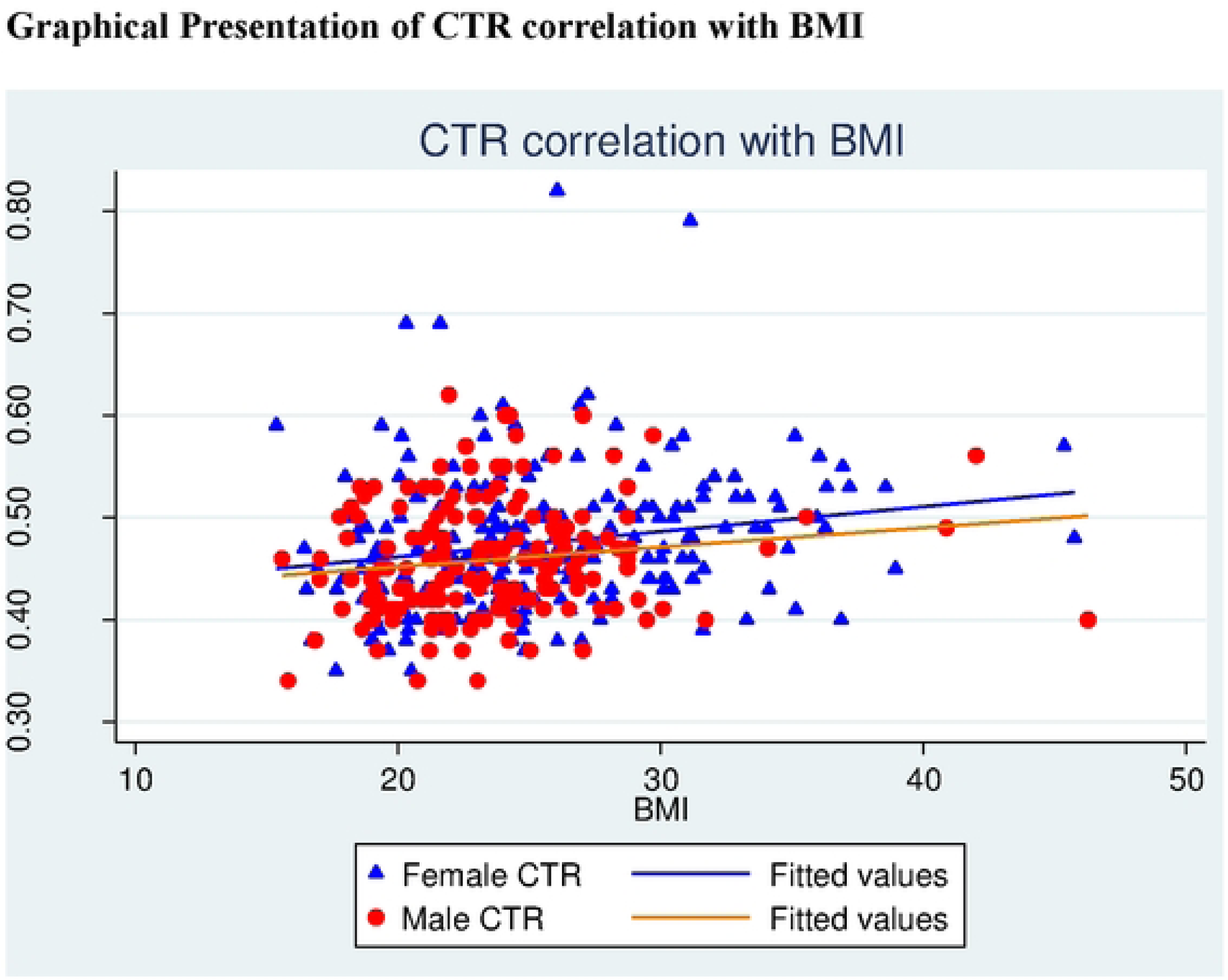
Correlation between strata ofBMI by gender with CTR.

**Fig 4.**
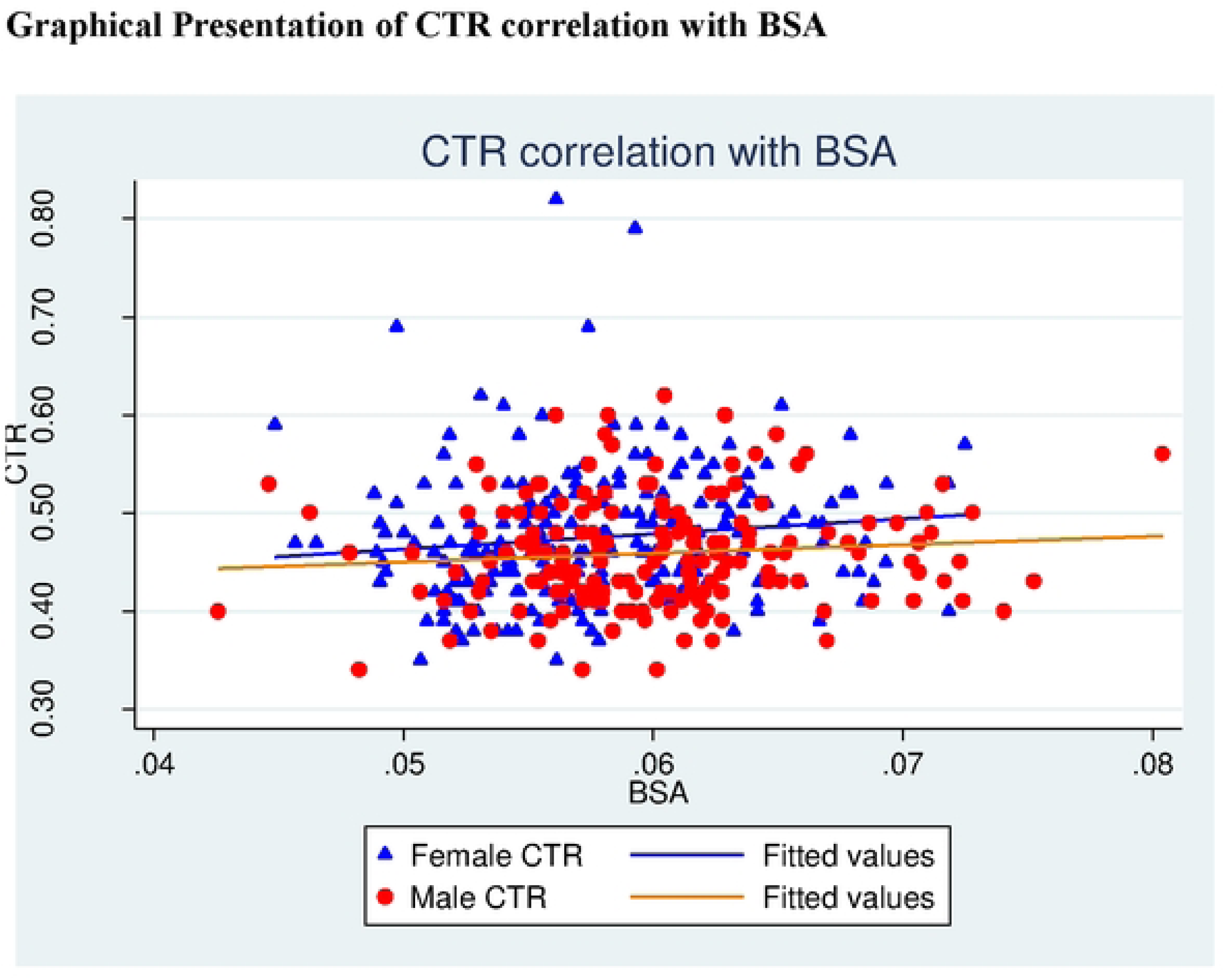
Correlation between strata of BSA by gender with CTR.

**Fig 5.**
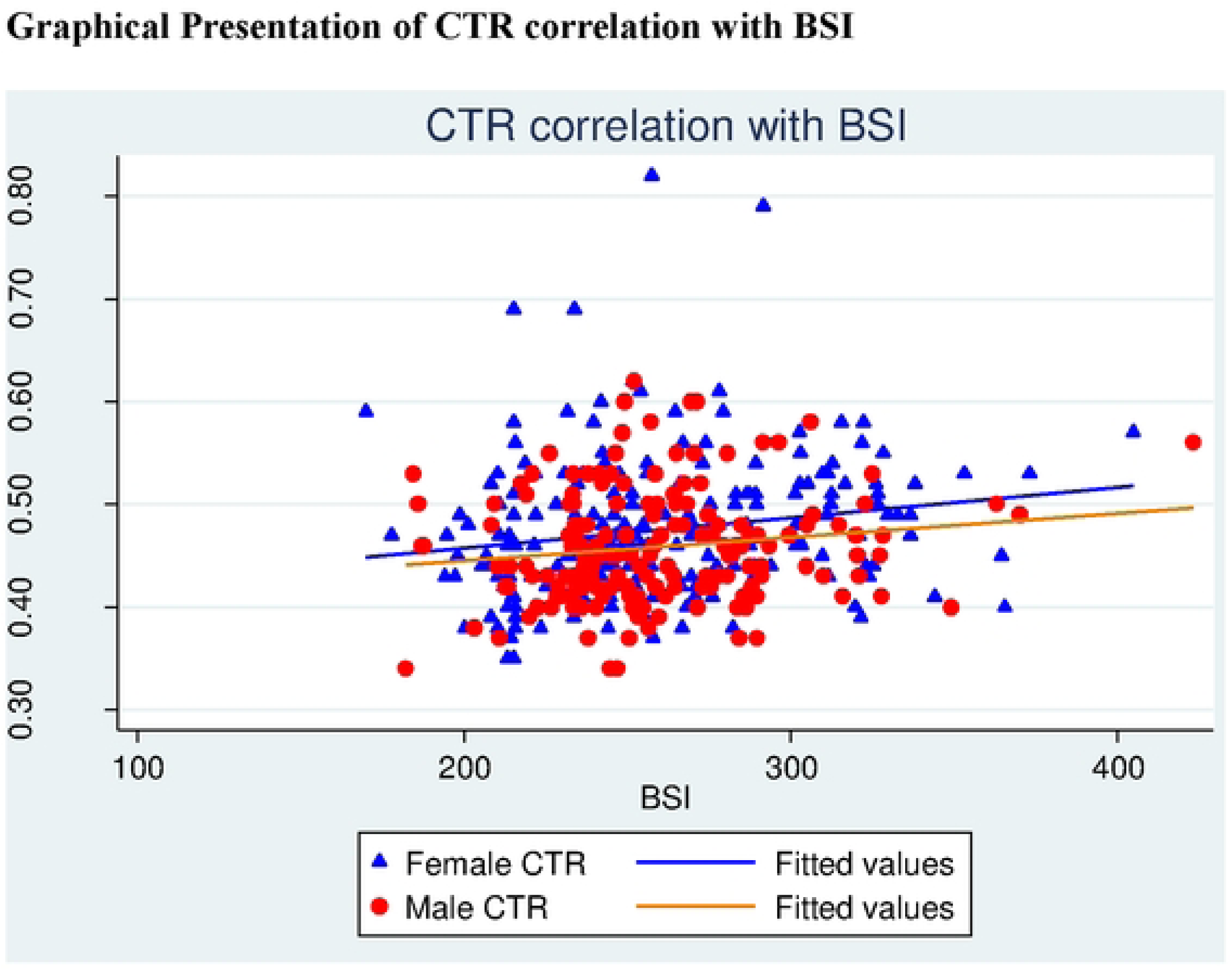
Correlation between strata of BSI by gender with CTR.

## REFERENCES

1. Mohammadi S, Hedjazi A, Sajjadian M, Ghoroubi N, Mohammadi M, Erfani SJJoc, et al. Study of the normal heart size in Northwest part of Iranian population: a cadaveric study. 2016;8(3):119.

2. McKee JL, Ferrier KJNMJ. Is cardiomegaly on chest radiograph representative of true cardiomegaly: a cross-sectional observational study comparing cardiac size on chest radiograph to that on echocardiography. 2017;130(1464):57–63.

3. Pfaffenberger S, Bartko P, Graf A, Pernicka E, Babayev J, Lolic E, et al. Size matters! Impact of age, sex, height, and weight on the normal heart size. 2013;6(6):1073–9.

4. Simkus P, Gutierrez Gimeno M, Banisauskaite A, Noreikaite J, McCreavy D, Penha D, et al. Limitations of cardiothoracic ratio derived from chest radiographs to predict real heart size: comparison with magnetic resonance imaging. 2021;12:1–10.

5. Truszkiewicz K, Poręba R, Gać PJJoCM. Radiological cardiothoracic ratio in evidence-based medicine. 2021;10(9):2016.

6. McKee J, Ferrier KJTNZMJ. Is cardiomegaly on chest radiograph representative of true cardiomegaly: a cross-sectional observational study comparing cardiac size on chest radiograph to that on echocardiography. 2017;130(1464):57–63.

7. Mensah Y, Mensah K, Asiamah S, Gbadamosi H, Idun E, Brakohiapa W, et al. Establishing the cardiothoracic ratio using chest radiographs in an Indigenous Ghanaian population: a simple tool for cardiomegaly screening. 2015;49(3):159–64.

8. Truszkiewicz K, Macek P, Poręba M, Poręba R, Gać PJRR, Practice. Radiological cardiothoracic ratio as a potential marker of left ventricular hypertrophy assessed by echocardiography. 2022;2022(1):4931945.

9. Ekedigwe JE, Pam SD, Binitie PO, Sirisena AU, Hameed M, Adegbe EOJJoMitT. Cardiothoracic ratio and body mass index in normal young adult Nigerians. 2014;16(2):47–51.

10. Emegoakor A, Ukoha UJIJoB, Applied, Research I. Relationship between cardiothoracic ratio and some selected anthropometric parameters in relation to gender in an adult Nigerian population. 2018;7(3):83–91.

11. Mehra S, Kumar S. Study of interrelationship between heart diameter and cardio-thoracic ratio with body habitus: a hospital based study to evaluate cardiac enlargement. 2019.

12. Sb CH, Browner W, Grady D, Newman T, Gaertner R. Designing clinical research: an epidemiologic approach. 2013.

13. Hattori K, Ishii T, Sato N, Motegi T, Yamada K, Gemma A, et al. Association Between Cardiothoracic Ratio On Plain Chest Roentgenogram And Cardiopulmonary Function In Patients With Chronic Obstructive Pulmonary Disease. B24 COPD AND CARDIOVASCULAR DISEASE: American Thoracic Society; 2011. p. A2622–A.

14. Mohajan D, Mohajan HKJJoIiMR. Body mass index (BMI) is a popular anthropometric tool to measure obesity among adults. 2023;2(4):25–33.

15. Hall. BFCA. Body Surface Area. National Library of Medicine. 2022.

16. Shirazu I, Sackey T, Tiburu EK, Mensah Y, Forson AJIJRSET. The use of body surface index as a better clinical health indicators compare to body mass index and body surface area for clinical application. 2018;4(11):131–6.

17. Msuega C, Hameed M, Annongu IJJMSCR. Establishing the cardiothoracic ratio of nonhypertensive patients using posteroanterior (PA) chest radiographs in an indigenous Nigerian population. 2021;9(2):1-9.

18. Ominde BS, Enakpoya P, Ikubor J, Ovovwero OJGIJHSr. Assessment of the cardiothoracic ratio and its association with gender and age: a Nigerian study. 2023;8(2):1622.

19. Brakohiapa EK, Botwe BO, Sarkodie BDJEJoHS. Gender and age differences in cardiac size parameters of ghanaian adults: Can one parameter fit all? part two. 2021;31(3).

20. Nimusiima E, Wafula ST, Mendoza H, Ndejjo R, Atusingwize EJIH. Physical activity levels and sociodemographic factors associated with meeting recommended levels among shop attendants in Mbarara municipality, Uganda. 2022;14(2):183–8.

21. Olander R, Sundholm J, Ojala T, Andersson S, Sarkola TJUiO, Gynecology. Differences in cardiac geometry in relation to body size among neonates with abnormal prenatal growth and body size at birth. 2020;56(6):864–71.

22. Shirazu I, Sackey TA, Tiburu EKJIJSRSET. Determination of Standard Reference Cardiothoracic Ratio and the Relationship with Body Parameters as A Patients Health Indicator for Clinical Application. 2019;6(5):318–226.

23. Halilu S, Aiyekomogbon J, Igashi J, Ahmed H, Aliyu YJAoIS. Cardiothoracic ratio on chest radiographs as a predictor of hypertensive heart disease among adults with systemic hypertension. 2017;7(3):82-.

24. Saheera S, Krishnamurthy PJCt. Cardiovascular changes associated with hypertensive heart disease and aging. 2020;29:0963689720920830.

25. Chou C-Y, Wang CC, Chiang H-Y, Huang C-F, Hsiao Y-L, Sun C-H, et al. Cardiothoracic ratio values and trajectories are associated with risk of requiring dialysis and mortality in chronic kidney disease. 2023;3(1):19.

26. Li Z, Hou Z, Chen C, Hao Z, An Y, Liang S, et al. Automatic cardiothoracic ratio calculation with deep learning. IEEE Access. 2019;7:37749–56.

27. Saiviroonporn P, Rodbangyang K, Tongdee T, Chaisangmongkon W, Yodprom P, Siriapisith T, et al. Cardiothoracic ratio measurement using artificial intelligence: observer and method validation studies. BMC Med Imaging. 2021;21(1):95.

